# Exhaustive white blood cell extraction and profiling across whole-slide images of blood and bone marrow smears

**DOI:** 10.64898/2026.10.23.26366335

**Authors:** Michael Deutges, Tabita Ghete, Altana Namsaraeva, Stephanie Sembill, Markus Metzler, Carsten Marr, Maximilian Buser

## Abstract

Blood and bone marrow smears are central to diagnosing hematological disorders. While whole-slide scanning enables automated single-cell profiling at scale, the computational burden of gigapixel images has confined automated analysis to manually pre-selected regions of interest. Here, we present HemAutomaton, a framework built on neural cellular automata that extracts white blood cells across entire whole-slide images without manual region selection and with minimal annotation requirements. HemAutomaton matches the in-domain detection accuracy of Faster R-CNN while surpassing it under domain shifts, operating at a fraction of the computational cost and with over 10,000-fold fewer parameters. Across three blood cancer cohorts, we extract over 100 million single-cell images and achieve annotation-efficient differential counting via few-shot classification using a pretrained hematology foundation model. Slide-wide spatial analysis uncovers consistent blast-frequency gradients across smears and demonstrates that automated blast counts match expert evaluations within inter-expert variability. Scaling automated profiling from hundreds to tens of thousands of cells per smear overcomes conventional sampling limits and establishes the foundation for more sensitive AI supported hematological assessments.

## Introduction

Peripheral blood (PB) and bone marrow (BM) smears serve as the primary first-line diagnostic modality for the identification and characterization of hematological malignancies and cytopenias. Diagnosis of these acute and chronic blood disorders—including myeloid and lymphoid leukemias, myelodysplastic syndromes, and myeloproliferative neoplasms—requires high-resolution, single-cell morphological assessment to differentiate specific lineages and evaluate maturation stages [1–4]. In clinical practice, hematopathologists analyze cell-type composition, identify aberrant cellular populations, and quantify blast frequencies to establish definitive diagnostic and prognostic markers [5, 6]. Crucially, a comprehensive manual review extends beyond the mere analysis of isolated single-cell images; it incorporates macroscopic and contextual slide-level indicators—such as specimen hemodilution (contamination of bone marrow aspirates with peripheral blood), preparation artifacts, and staining quality—to validate the clinical integrity of the specimen before specific regions are selected for differential counting [7, 8].

While a single smear contains up to hundreds of thousands of evaluable single white blood cells [9], conventional manual workflows utilize only a minute fraction of this data. To maintain operational throughput despite the time constraints of manual microscopy analysis, guidelines standardize manual blood count differentials to a limited sample size of up to 200 cells [7, 10, 11]. This introduces substantial statistical variance, causing high sampling error for rare cell populations and inherently limiting the detection of minimal residual disease. Furthermore, because manual inspection relies on subjectively selected fields of view, the comprehensive characterization of the cellular composition and macroscopic spatial organization across the entire smear remains impractical. Routine digitization of smears into whole-slide images (WSIs) offers a path to overcome these limitations by enabling computational processing at the slide-wide scale [12, 13].

To automate smear assessment, deep learning methods have recently extended hematopathological analysis from manually selected regions of interest (ROIs) to WSIs by introducing automated pipelines for cell extraction [14]. Extracted single-cell images form the foundation for downstream analyses, including automated differential counting [9, 15–20], disease prediction [21–24], and vision-language modeling [25]. Most existing extraction pipelines mirror the manual clinical workflow, passing WSIs through sequential, specialized stages: ROI (pre)-selection, quality assessment, and high-resolution single-cell detection and segmentation.

These approaches, however, present several limitations: First, supervised cell detection typically relies on extensive expert annotations, such as manually bounded ROIs [9, 16, 26], single-cell bounding boxes [9, 16, 18, 24, 27], or pixel-level segmentation masks [21], which severely restricts training data scale and diversity. Second, cell detection is typically performed at full resolution using deep-learning architectures such as Faster R-CNN [9], YOLO variants [16, 18, 19, 24, 28], U-Net approaches [21], large vision foundation models such as SAM [20, 25], or other CNN-based architectures [26, 27], requiring substantial computational resources when processing entire WSIs. Third, external validation is often restricted to downstream task performance, such as differential counts or disease classification, rather than analyzing the underlying single-cell extraction pipeline itself [19–21]. Heavily parameterized networks trained on small, single-center cohorts tend to overfit to local staining profiles and scanner optics, making them vulnerable to performance degradation under domain shifts that are always present in real-world data from laboratories and clinics.

Beyond diagnosis, large-scale single-cell extraction is critical for training self-supervised hematology foundation models, such as DinoBloom, which learned morphological features from 380,000 curated white blood cell images [29]. By offering unified, transferable representations without task-specific retraining, learned single-cell embeddings form the foundation for diverse downstream tasks, including single-cell classification [30–32] and disease prediction [33–38]. However, scaling these architectures to millions of cells across heterogeneous cohorts remains constrained by the inability of existing extraction pipelines to adapt to varying staining protocols and scanning hardware. Limited dataset diversity causes self-supervised methods to focus on institutional signatures rather than biologically relevant morphological features. Efficient, slide-wide single-cell extraction is thus a key prerequisite for assembling the high-volume datasets required to advance the generalizability of future hematology foundation models.

To address these challenges, a whole-slide extraction framework must satisfy the following three criteria: (i) computational efficiency at the gigapixel scale, (ii) annotation efficiency to eliminate site-specific expert relabeling, and (iii) robustness against varying staining protocols, scanner hardware, and sample preparation. Neural cellular automata (NCAs) are uniquely suited to meet these constraints [39]. Originally introduced as a bio-inspired architecture for image generation through local cell-to-cell interactions [40], NCAs have recently demonstrated competitive performance in medical image segmentation, requiring substantially fewer trainable parameters than conventional deep learning models [41]. In hematology, NCAs have been applied to single-cell classification [42–44] and weakly supervised white blood cell segmentation [45], demonstrating strong robustness under cross-domain evaluation.

Here, we present HemAutomaton, a lightweight pipeline for large-scale white blood cell extraction directly from WSIs. Built upon a parameter-efficient NCA model optimized for cell localization, HemAutomaton operates on the premise that cell identification relies on local context rather than long-range features, enabling high-throughput detection at the full-slide scale. By processing the complete slides without requiring ROI pre-selection, the pipeline eliminates the need for manual intervention. We demonstrate that HemAutomaton enables the extraction of 108 million single-cell images across three independent clinical cohorts with minimal expert supervision. Using the extracted cells, we further demonstrate its utility in two downstream applications: automated slide-level differential cell counting and quantification of macroscopic spatial blast gradients across entire smears.

## Results

### HemAutomaton whole-slide single-cell extraction pipeline

HemAutomaton implements a multi-stage pipeline for scalable white blood cell extraction across gigapixel WSIs, without manual region selection (Figure 1). We exemplified the framework using three independent datasets spanning BM and PB smears from five diseases and normal controls, acquired on four scanning systems (Figure 1A and Methods): (i) an in-house B-cell precursor acute lymphoblastic leukemia (B-ALL) dataset containing paired BM and PB WSIs from 120 patients; (ii) a BM smear dataset released by Höfener et al. [18] comprising 246 patients diagnosed with acute lymphoblastic leukemia (n = 100), acute myeloid leukemia (n = 95), and chronic myeloid leukemia (n = 51); and (iii) a publicly available PB dataset released by de Almeida et al. [21] containing 362 cropped WSIs from individuals diagnosed with myelodysplastic syndromes (n = 233), anemias (n = 71), and hematologically normal controls (n = 58). WSIs from different datasets exhibit substantial heterogeneity arising from differences in specimen type (BM versus PB), laboratory workflows, cell density, staining intensity, and preparation artifacts, illustrated by representative whole-slide views and magnified regions. An overview of the available annotations for each dataset and the usage throughout the study is provided in Supplementary Figure 1. Because diagnosis of the disorders in these datasets relies on white blood cell rather than red blood cell morphology, we focused exclusively on leukocyte detection and analysis.

**Figure 1:**
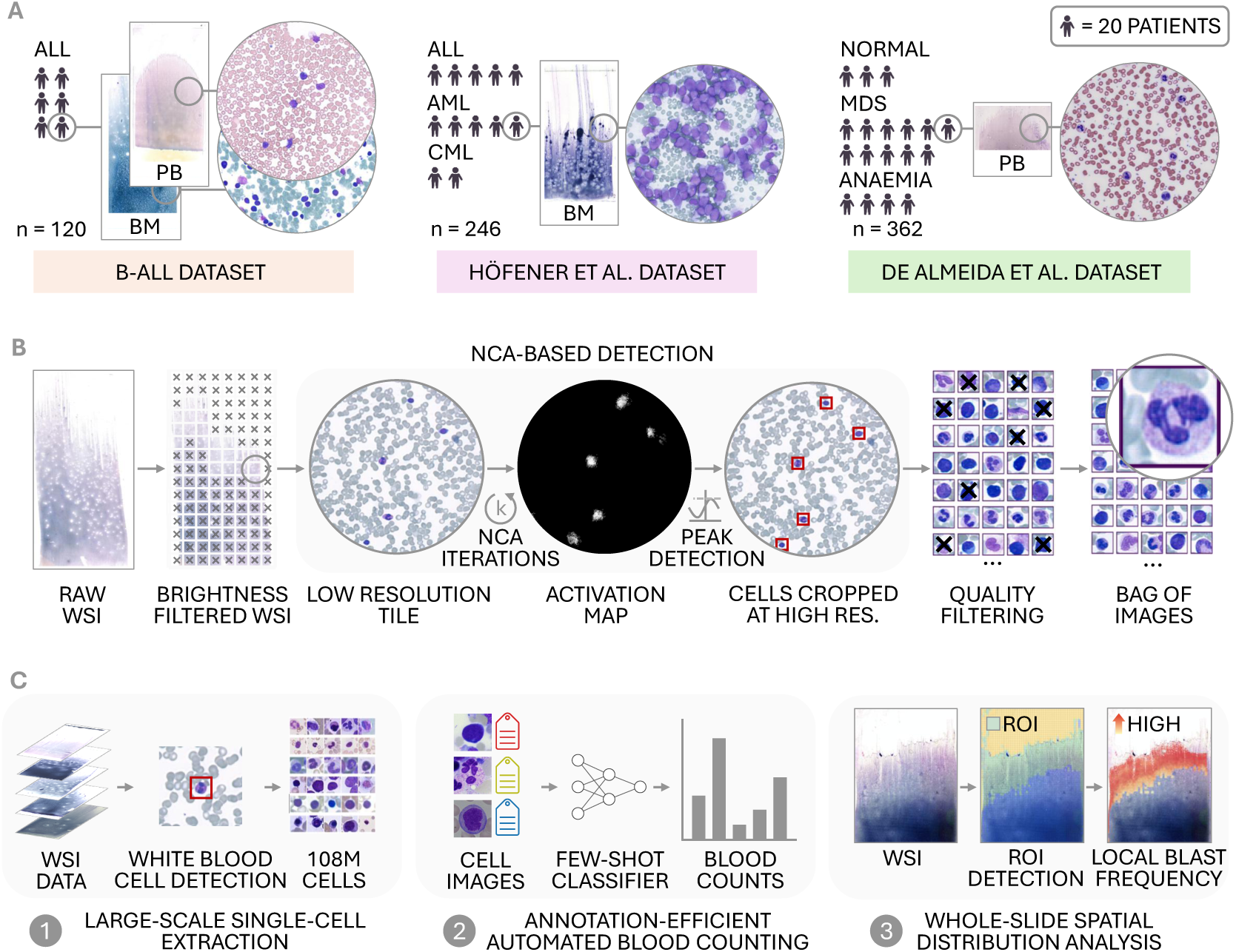
HemAutomaton enables exhaustive white blood cell extraction and slide-wide analysis of hematology smears. **(A)** We demonstrate WSI smear data processing on three independent cohorts: (i) a B-cell precursor acute lymphoblastic leukemia dataset (B-ALL) with paired BM and PB samples from 120 individuals; (ii) a BM smear dataset released by Höfener et al. [18] comprising slides from 246 individuals across three leukemia subtypes, including acute lymphoblastic leukemia (ALL, n = 100), acute myeloid leukemia (AML, n = 95), and chronic myeloid leukemia (CML, n = 51); (iii) 362 PB smears released by de Almeida et al. [21] from individuals with myelodysplastic syndromes (MDS, n = 233) and anemias (n = 71), as well as hematologically normal controls (n = 58). **(B)** HemAutomaton first partitions raw WSIs into tiles and filters out uninformative background and hyper-dense cell aggregates using brightness heuristics. Next, single-cell coordinates are identified using an NCA-based model applied to WSI tiles rescaled to a standardized resolution of approximately 1 µm per pixel (Methods). The NCA is trained to efficiently identify white blood cell locations, producing smooth, cell-shaped activation blobs with a high-intensity peak at the cell centers, which encourages consistent localization across heterogeneous smear conditions. Single-cell coordinates are detected as local maxima of the resulting activation maps. These are mapped back to the original full-resolution WSI, and high-resolution single-cell patches are extracted at the maximum available resolution. A quality-filtering model discards low-quality single-cell patches, such as smudged or blurry cells, resulting in high-quality single-cell images. **(C)** We demonstrate the utility of HemAutomaton across three applications: (1) extracting over 100 million single-cell images from PB and BM smear WSIs as a resource for future representation learning; (2) performing data-efficient single-cell classification for automated differential counts by combining pretrained morphological embeddings [29] with sparse target-domain annotations; and (3) mapping classified cells back to their spatial coordinates to quantify cellular distributions and tissue heterogeneity across entire slides.

In the first processing stage, raw WSIs are partitioned into non-overlapping tiles, followed by a brightness-based filter that discards uninformative background regions and densely aggregated cell areas where individual cells cannot be resolved (Figure 1B). Next, to optimize computational efficiency during slide-wide inference, the remaining tiles are downsampled to a standardized resolution and processed using an NCA-based detection model that iteratively generates cell-shaped activation maps with peaks at white blood cell centers. These activation maps are smoothed and converted into cell center coordinates via local-maxima detection, which are then mapped back onto the original full-resolution WSIs for the extraction of single-cell patches. Finally, an automated quality-filtering model discards blurred, damaged, or otherwise unsuitable crops, yielding a collection of high-quality single-cell images for downstream analyses (Methods).

We validated HemAutomaton’s utility for large-scale dataset generation, automated differential cell counting, and spatial analysis of blast distributions (Figure 1C).

### Robust and scalable white blood cell detection

Accurate white blood cell localization across heterogeneous preparation and scanning protocols is essential for automated whole-slide analysis, given the variability across clinical laboratories and within individual slides. To enable efficient whole-slide processing, we performed detection at a reduced standardized resolution of approximately 1 µm per pixel and benchmarked our NCA-based detection model against Faster R-CNN [9, 46] and a simple blob detection baseline (Methods) based on color and intensity (Table 1). We evaluated this localization task in a three-domain cross-validation setup (Methods).

**Table 1:** NCA-based white blood cell detection generalizes across domains at reduced computational cost. ROI-level F1 scores for cell detection across data from the B-ALL, Höfener et al., and de Almeida et al. datasets are reported for HemAutomaton’s NCA-based detection model, Faster R-CNN, and a blob detection baseline. For each source-target combination, e.g., train on B-ALL, test on Höfener et al., performance was estimated using five-fold cross-validation. Performance scores are presented as mean ± standard deviation across three in-domain settings and six cross-domain evaluations for each method. The F1 Gap denotes the mean performance drop between in-domain and cross-domain evaluations, with lower values indicating greater robustness to domain shift. While Faster R-CNN achieved the highest mean in-domain performance, the NCA-based detection model showed the strongest mean cross-domain performance and the smallest generalization gap. Additionally, NCA-based detection substantially reduces computational cost compared to Faster R-CNN. Efficiency is assessed in terms of model size, computational demand, inference speed, and memory consumption during tile-based detection on WSIs using 512 × 512 tiles. The number of trainable parameters characterizes model size, while FLOPs quantify the number of computational operations required per tile. Latency and throughput capture inference speed, and peak RAM and VRAM quantify the maximum memory required during inference (Methods). The blob baseline shows high efficiency due to its minimal computational requirements as a simple threshold-based baseline. However, it does not provide competitive detection performance under realistic conditions. Compared to Faster R-CNN, the proposed NCA-based detection model requires over 10,000-fold fewer parameters and is more efficient across all other metrics.

| Metric | Blob | Faster R-CNN | NCA |
| --- | --- | --- | --- |
| *Detection performance* |  |  |  |
| F1 In-domain (mean) ↑ | $0.59 \pm 0.16$ | **$0.87 \pm 0.02$** | $0.85 \pm 0.01$ |
| F1 Cross-domain (mean) ↑ | $0.51 \pm 0.18$ | $0.65 \pm 0.18$ | **$0.79 \pm 0.11$** |
| F1 Gap (mean) ↓ | $0.08 \pm 0.26$ | $0.21 \pm 0.14$ | **$0.06 \pm 0.10$** |
| *Computational efficiency* |  |  |  |
| Params (trainable) ↓ | 0 | 41.4M | 3.3k |
| FLOPs (per tile) ↓ | 26.2M | 126.1G | 10.0G |
| Latency ↓ | 14.6 ms | 32.4 ms | 17.7 ms |
| Throughput ↑ | 69.1 tiles/s | 30.9 tiles/s | 56.3 tiles/s |
| Peak RAM ↓ | 1.4 GB | 1.4 GB | 1.0 GB |
| Peak VRAM ↓ | 0 GB | 0.6 GB | 0.3 GB |

In domain, Faster R-CNN achieved the highest average ROI-level F1 score (0.87 ± 0.02; mean ± standard deviation over n = 3 in-domain settings), with HemAutomaton’s NCA detection model performing competitively (0.85 ± 0.01), whereas the blob baseline showed substantially lower overall performance (0.59 ± 0.16). Under domain shift, i.e., when the model is trained on one dataset and applied to another, the NCA model achieved the highest average cross-domain performance (F1 score of 0.79 ± 0.11 across n = 6 cross-domain settings), outperforming Faster R-CNN (0.65 ± 0.18) and the blob baseline (0.51 ± 0.18). Consistent with this, the NCA model showed the smallest average gap between in-domain and cross-domain F1 scores (0.06 ± 0.10), indicating more stable performance across datasets than Faster R-CNN (0.21 ± 0.14) and the blob baseline (0.08 ± 0.26). Notably, the relatively small gap of the blob baseline reflected uniformly poor absolute performance rather than strong transferability. Together, these results indicate that the proposed NCA-based detection model achieves strong cross-domain generalization while maintaining competitive in-domain performance. The full experiment matrix resolving all source-target combinations is provided in Supplementary Table 1.

In addition to detection performance, we evaluated the computational efficiency of the proposed approach and compared it to Faster R-CNN and the blob baseline (Table 1). Efficient processing is critical for practical deployment in laboratories and clinics, where large numbers of images must be analyzed every day. The NCA-based detection model requires four orders of magnitude fewer parameters than Faster R-CNN (3.3k versus 41.4M) and reduces computational cost in terms of FLOPs per tile by a factor of 12.6 (10.0G versus 126.1G). This translates into 0.55-fold lower latency and 1.8-fold higher throughput during inference, demonstrating substantially faster processing compared to Faster R-CNN, while maintaining competitive detection performance. The NCA-based approach of HemAutomaton also minimizes GPU memory footprint during inference, facilitating slide-wide analysis on resource-constrained hardware. While the blob baseline exhibits excellent computational efficiency, its low localization accuracy (Table 1) limits applicability.

Together, these results show that the NCA-based detection model matches Faster R-CNN’s in-domain performance and exceeds it under domain shift at reduced computational cost.

We next assessed annotation efficiency by evaluating Faster R-CNN versus our NCA-based detection model across label-scarce settings and under domain shifts (Figure 2A). The blob baseline was excluded from this experiment due to its limited performance in the full data benchmark (Table 1). WSIs exhibit substantial inter- and intra-dataset variability (Figure 1A), including differences in cell density, staining intensity, background composition, and imaging artifacts. This variability requires models to generalize from limited and potentially non-representative training data.

**Figure 2:**
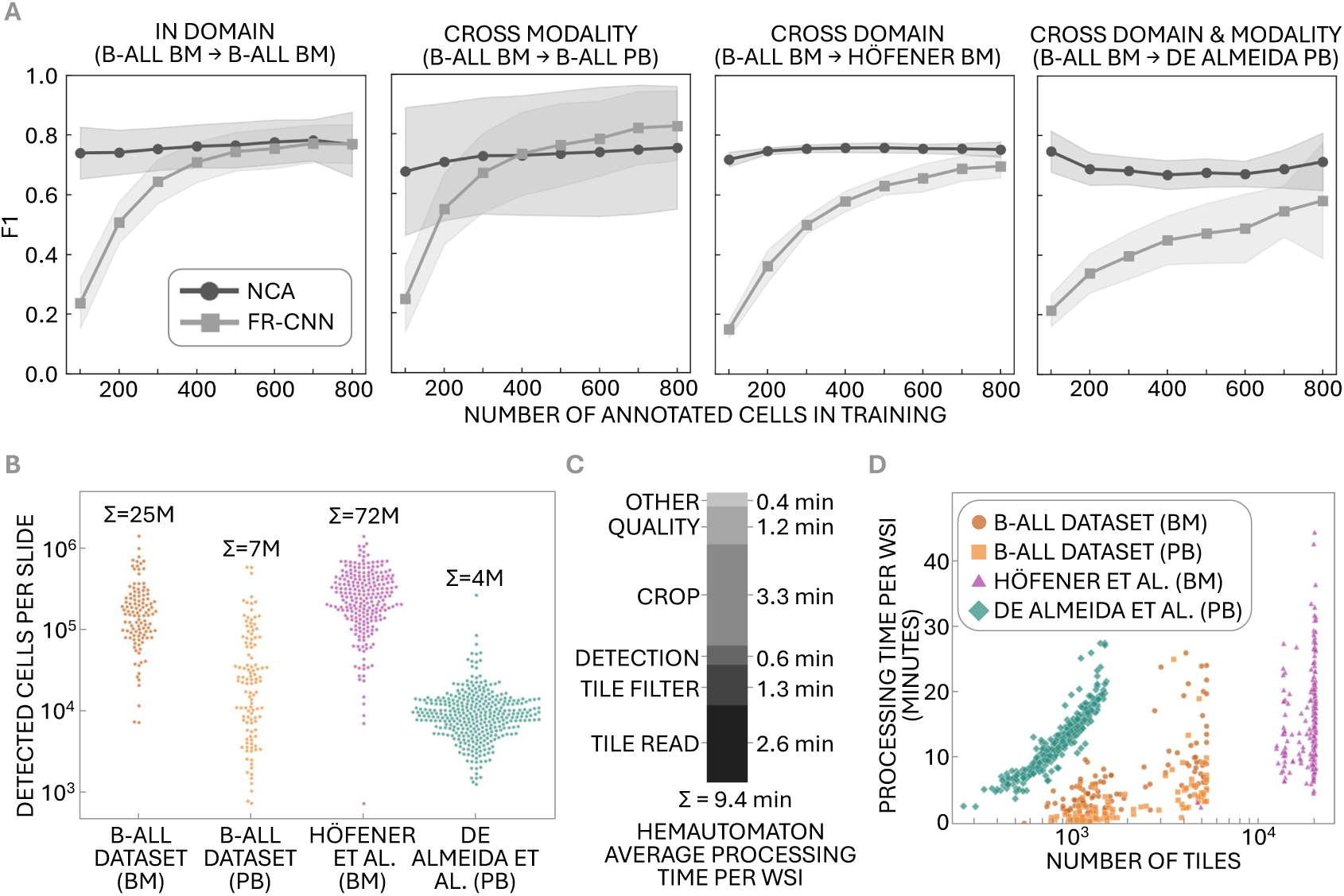
HemAutomaton provides robust and scalable white blood cell detection across heterogeneous whole-slide images. **(A)** NCA-based detection remains robust with few annotated cells, even under domain shifts and across different modalities. It outperforms Faster R-CNN in the extreme low-annotation regime across all evaluation settings and requires as little as a single training patch (100 annotated cells). Lines and shaded areas indicate mean F1 scores and standard deviations across five validation folds, respectively. **(B)** Automated single-cell extraction scales to millions of cells while maintaining efficient whole-slide processing. In total, the pipeline extracts over 100 million white blood cell images, with Σ *=* 25*M* single-cell patches from BM (*n =* 120 slides) and Σ *=* 7*M* from PB (*n =* 120 slides) within the B-ALL dataset, as well as Σ *=* 72*M* from the Höfener et al. BM dataset (*n =* 246 slides), and Σ *=* 4*M* from the de Almeida et al. PB dataset (*n =* 362 slides). **(C)** Processing a complete WSI takes less than ten minutes on average, with the runtime breakdown showing the contribution of tile reading, tile filtering, NCA-based detection, cell cropping, quality control, and other processing steps. **(D)** Across individual slides, processing time generally increases with the number of processed tiles, while systematic differences between datasets exist, reflecting dataset-specific characteristics such as image format and cell density.

Trained and tested in-domain on B-ALL BM samples, Faster R-CNN exhibits steady performance gains as more training data becomes available (Figure 2A). In contrast, the NCA-based detection model achieves its near-maximum performance with as little as 100 white blood cells contained in a single training patch. A similar pattern is observed under domain shifts: When trained on B-ALL BM smears and evaluated on B-ALL PB data and the independent Höfener et al. and de Almeida et al. datasets, Faster R-CNN shows substantially stronger dependence on training set size, with severe degradation in the low-data regime of 100 to 800 annotated cells. By contrast, the NCA-based detection model maintains robust performance across all evaluation settings (B-ALL BM *→* B-ALL PB, B-ALL BM *→* Höfener BM, B-ALL BM *→* de Almeida PB), with the clearest advantage observed for the de Almeida et al. dataset, which represents the most challenging validation scenario as it includes both cross-modality (BM versus PB) and cross-institution domain shifts.

Achieving near-maximum performance from a single 100-cell training patch demonstrates the NCA-based detection model’s annotation efficiency, enabling direct model transfer to new datasets with minimal manual labeling effort while mitigating performance degradation on unseen or heterogeneous smear regions.

Having established robustness, computational efficiency, and annotation efficiency of the NCA-based detection model, we used HemAutomaton to automatically extract millions of white blood cells from entire WSIs across three distinct datasets (Figure 2B). The pipeline detects an average of 1.3 *×* 10^5^ white blood cells per slide (B-ALL BM: 220*k ±* 210*k*, mean *±* standard deviation, *n =* 120 slides; B-ALL PB: 57*k ±* 99*k*, *n =* 120; Höfener et al.: 300*k ±* 220*k*, *n =* 246; de Almeida et al.: 11*k ±* 16*k*, *n =* 362), totaling 108*M* single-cell patches from BM and PB slides and providing a large-scale unlabeled resource of single-cell images.

Despite this substantial data scale, whole-slide processing remains tractable (Figure 2C). Processing an entire WSI takes approximately 9.4 minutes on average, of which NCA-based detection accounts for only approximately 0.6 minutes. Tile reading and high-resolution cell cropping account for the largest contributions to overall processing time, while tile filtering, quality control, and other processing steps contribute comparatively little additional runtime (for a detailed breakdown see Supplementary Figure 2). Processing times vary across datasets, reflecting differences in image dimensions, cellular content, and data formats (Figure 2D).

We compared HemAutomaton with 12 published white blood cell detection pipelines [9, 15–25]. Only 3 out of 12 provided publicly available implementations and pretrained models (Supplementary Table 2). In the ROI-level benchmark across all three datasets, HemAutomaton achieved the highest average F1 score (0.79, Supplementary Table 3). We additionally applied two available end-to-end pipelines, one from Tayebi et al. [16] and Haemorasis [21] to representative WSIs without modification (Supplementary Figure 3), uncovering transferability and deployment bottlenecks. Neither pipeline supported DICOM formats, precluding evaluation on the Höfener et al. dataset. On supported MRXS and TIFF formats, Tayebi et al. discarded *>* 99% of tiles during heuristic preprocessing, resulting in severe under-detection (*<* 150 cells per slide). While Haemorasis detected more candidates, it required substantially longer processing times than HemAutomaton and frequently extracted non-WBC artifacts, whereas HemAutomaton achieved rapid full-slide inference without excessive tile rejection or loss of specificity.

### Annotation-efficient automated blood counts

Extracted single-cell images form the basis for slide-wide morphological analyses, such as differential blood counts. However, cell classification typically requires extensive expert annotation. Pretrained hematology foundation models can reduce these annotation requirements by providing transferable morphological representations learned from large collections of single-cell images. To minimize expert annotation effort, we therefore used DinoBloom [29] as a frozen feature extractor and investigated whether sparse supervision suffices for cell-type classification (Figure 3A).

**Figure 3:**
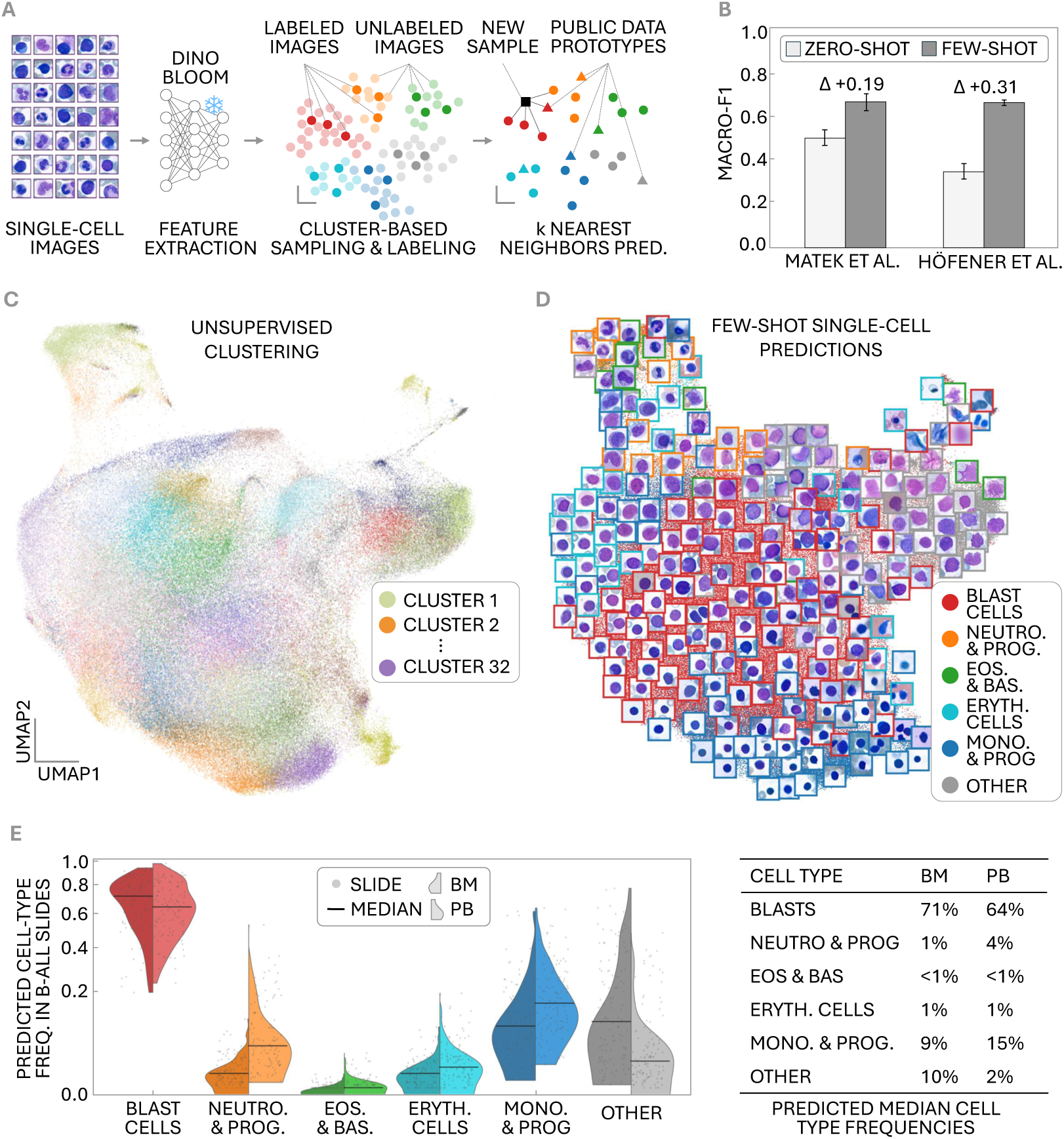
Few-shot single-cell classification enables labeling-efficient automated blood counts. **(A)** Few-shot learning via labeled single-cell images enables efficient annotation of target-domain data. Single-cell images are embedded using a frozen feature extractor [29], and representative samples are selected via unsupervised clustering for manual labeling. The resulting labeled cells are used for k nearest-neighbor classification of new samples. To account for underrepresented or missing cell types in the few-shot set, public data prototypes (mean feature vectors of all available cells within each class) are incorporated and reweighted based on the availability of the corresponding cell type in the target-domain annotations. **(B)** Few-shot supervision improves classification performance compared to zero-shot transfer. Labeling n = 20 target-domain samples per cluster increases F1 scores by 0.19 (from 0.48 to 0.67) and 0.31 (from 0.37 to 0.67), respectively, relative to the zero-shot setting. **(C)** UMAP projection of single-cell embeddings obtained from a DinoBloom model shows a structured feature space underlying the few-shot classification approach with k-means clustering results used for sample selection in the few-shot annotation workflow. **(D)** Nearest-neighbor predictions across the embedding space show morphologically meaningful organization of cell types and artifacts. For qualitative assessment of the classification results, randomly selected single-cell images are overlaid at their corresponding embedding positions, with frame colors indicating cell-type predictions. **(E)** Few-shot classification enables automated estimation of blood counts per slide. Across 120 B-ALL slides with 1000 sampled single cells each, distributions are plausible, with elevated blast frequencies reflecting the disease and reduced mature hematopoietic populations in BM (left) as compared to PB (right). Cells predicted as “OTHER” mainly contain smudged cells and artefacts, which are typically not considered in manual blood counts.

To maximize annotation efficiency, representative cells were selected by clustering the target-domain embeddings using k-means. Annotating a fixed number of cells per cluster ensures coverage across the full morphological spectrum, avoiding bias toward high-frequency cell populations. To compensate for cell types that are absent or only sparsely represented in few-shot reference training data, we additionally incorporated class prototypes derived from publicly available labeled datasets [18, 31] with a custom weighting scheme. Cell-type prediction for the remaining cells was performed using a nearest-neighbor classifier (Methods).

We first evaluated the few-shot approach on the Matek et al. [31] and the Höfener et al. [18] single-cell datasets (Figure 3B). To establish a common label space across datasets, annotations from the two datasets were harmonized using a common set of cell types including blast cells, neutrophils and neutrophil progenitors, eosinophils and basophils, nucleated erythroid cells, mononuclear cells and progenitors, and other cells and artifacts (Supplementary Figure 4). As a zero-shot baseline, cells from each dataset were classified using only labeled cells from the respective other dataset. Incorporating limited target-domain supervision improves classification performance over the zero-shot baseline (Figure 3B). Annotating only 20 cells per cluster increases macro-F1 scores by 0.19 (from 0.48 to 0.67) on the Matek et al. dataset and by 0.31 (from 0.37 to 0.67) on the Höfener et al. dataset, demonstrating that high classification accuracy can be achieved with minimal annotation effort. The full confusion matrices and results from an ablation experiment for varying numbers of labeled single-cell images per cluster are provided in the Supplementary Material (Supplementary Figure 5, Supplementary Table 4).

We next applied the few-shot workflow to automatically extracted cells from PB and BM WSIs of the B-ALL dataset. The resulting single-cell image features form a morphology landscape in which visually similar cell types cluster together, providing a basis for nearest-neighbor cell-type classification (Figure 3C). Applying the classifier to all extracted cells enables automated estimation of slide-level cell-type frequencies. Qualitative inspection of the predictions via UMAP revealed coherent regions of predicted cell types within the morphology landscape (Figure 3D). The resulting distributions are biologically plausible with elevated blast frequencies reflecting the disease, while PB samples exhibit relatively higher proportions of mature hematopoietic cells than the corresponding BM samples (Figure 3E).

### Spatial organization of blast cells

Automatically extracted and classified cells enable quantitative analysis of cellular distributions across complete WSIs (Figure 4A). While manual differential counts are performed in selected smear regions and evaluate only a limited number of cells, automated whole-slide analysis provides the opportunity to systematically investigate how cell-type composition varies across the entire specimen. As blast frequency is a key metric in leukemia diagnostics, we focused our spatial analysis on the distribution of blast cells. To ensure high-confidence blast identification, we adapted the few-shot classification approach to a binary blast versus non-blast classification, using 4, 000 expert-annotated cells from BM and PB smears.

**Figure 4:**
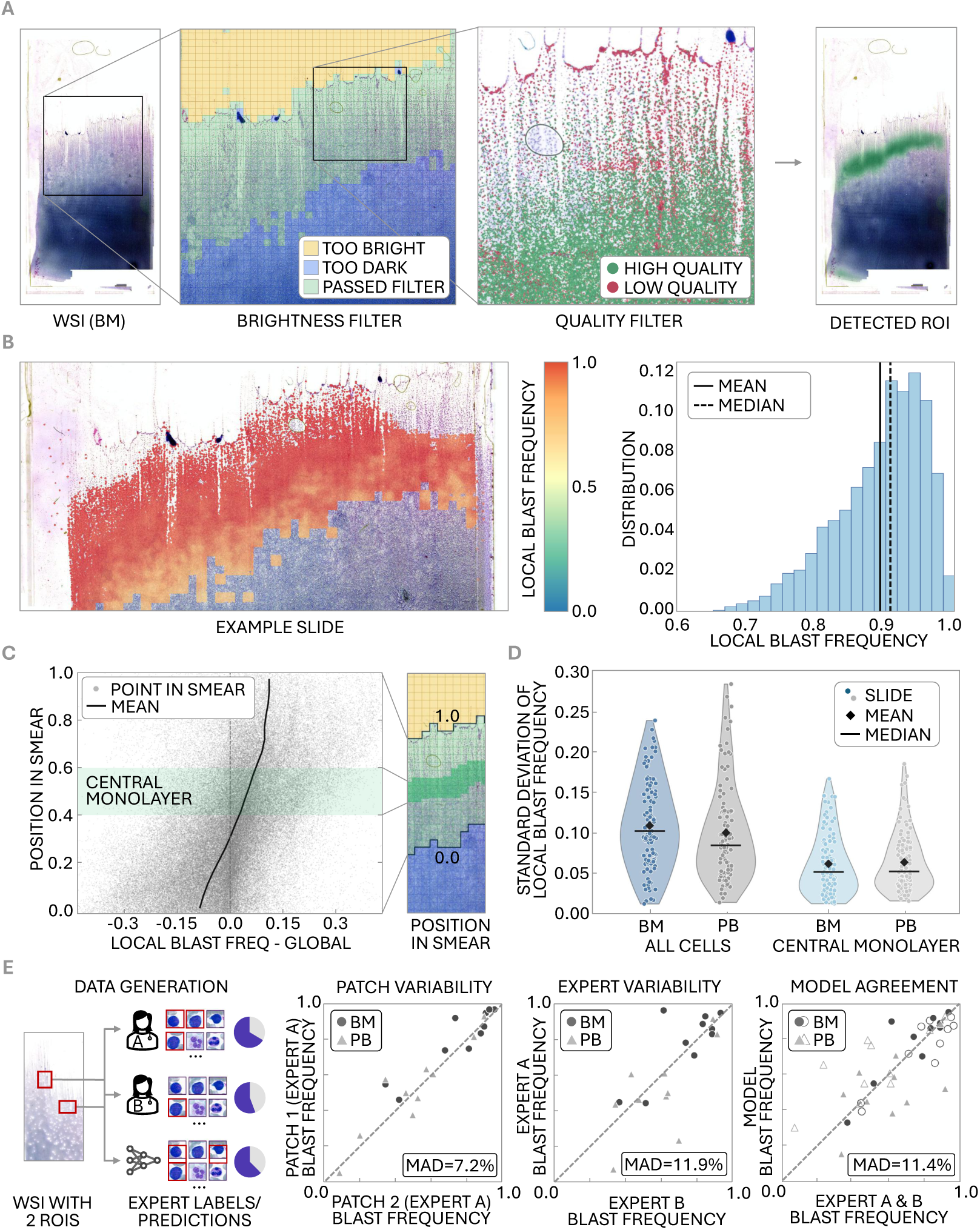
Whole-slide analysis reveals spatial heterogeneity of blast distributions and allows for expert-level blast frequency estimation. **(A)**Automated ROI detection identifies smear regions with analyzable single cells. WSI tiles are first filtered based on brightness to exclude background and overly dense regions containing overlapping cells. Single cells are subsequently detected in the remaining tiles and a quality filter is applied to remove low-quality images and artifacts. The resulting ROI defines the area considered during downstream spatial analysis. **(B)** Local blast frequencies can vary substantially within individual slides. Blast frequencies are estimated in local neighborhoods and visualized as a spatial heatmap for a representative BM slide. The analysis is restricted to the automatically identified analyzable smear region. The corresponding histogram shows the distribution of local blast frequencies across all neighborhoods. Local blast frequencies range from approximately 0.5 to 1.0, indicating pronounced spatial heterogeneity within a single smear. **(C)** Local blast frequencies show a systematic spatial gradient, with higher frequencies near the smear edge (position in smear = 1) and lower frequencies towards the center (position in smear = 0). The plot shows deviation of local blast frequency from the slide-level blast frequency along the smear direction **(D)**Spatial variability in blast frequencies is reduced when focusing on a strictly defined monolayer region. The distribution of standard deviations of local blast frequencies across slides for BM and PB samples is compared between all analyzable cells and those within the central 20% (positions 0.4–0.6) of the analyzable smear region, used as an approximation of the monolayer. **(E)** Validation of local blast-frequency estimates across 20 B-ALL slides (10 BM and 10 PB). Two independent experts (A and B) annotated the same 100 cells in each of two manually selected monolayer regions per slide. Scatterplots show patch-to-patch variability based on expert A annotations (MAD = 7.2%), inter-expert agreement across slides (aggregated patches; MAD = 11.9%), and agreement between model-derived and expert-derived blast frequencies (aggregated patches; cross-model MAD = 11.4%). Model predictions were obtained using patient-level cross-validation, with separate classifiers trained on annotations from either expert and evaluated against the respective other expert on held-out patients. Filled markers correspond to the model trained on expert A annotations and evaluated on expert B annotations, while empty markers correspond to the reverse setup. Non-inferiority of model-expert agreement relative to inter-expert agreement was demonstrated using the mean difference in absolute errors across the two model evaluations in a one-sided Wilcoxon signed-rank test (*n =* 20; margin = 5%; *p =* 0.0032).

Whole-slide cell-type prediction reveals pronounced spatial heterogeneity in blast frequencies across different regions of the same slide (Figure 4B). To quantify these effects across the B-ALL dataset, we analyzed local blast frequencies as a function of smear position (Figure 4C). A consistent gradient is observed, with elevated blast frequencies near the feathered edge (normalized smear position 0.9*−*1.0; on average 11.5% above the global mean) and progressively lower frequencies towards the denser central regions of the smear (normalized smear position 0 *−* 0.1; on average 9.4% below the global mean). This observation is consistent with established smear preparation effects: Differences in cell size and mechanical properties may lead to systematic enrichment or depletion of specific cell types in different regions [47]. Our observations therefore provide a quantitative rationale for the established clinical practice of restricting manual differential counts to particular monolayer regions while avoiding the feathered edge and densely aggregated areas. Restricting the analysis to a strict monolayer region substantially reduces the observed variability. Nevertheless, measurable spatial heterogeneity persists even within this region, with mean standard deviations of 5.8% and 6.0% in BM and PB, respectively, indicating that representative monolayer sampling reduces, but does not completely eliminate local variation in blast frequencies (Figure 4D).

To validate the automated blast-frequency estimates, we conducted an independent evaluation with a second expert across 20 B-ALL slides (10 patients with one PB and one BM slide each; Figure 4E). For each slide, two ROIs containing 100 cells each were manually selected from suitable monolayer regions and independently annotated by both experts (A and B), yielding 4, 000 cells annotated by both experts. Blast frequencies varied between the two ROIs (mean absolute difference MAD = 7.2%, based on expert A annotations). Because ROI selection was not standardized along the smear axis, these differences cannot be directly linked to the systematic spatial gradient characterized above. Moreover, given the limited number of cells per ROI, they may reflect both sampling variance and residual heterogeneity within the monolayer.

To evaluate automated blast-frequency estimation, we retrained two binary classifiers using annotations from either expert and evaluated their predictions against the respective independent expert using 10-fold patient-level cross-validation. Model-derived blast frequencies (aggregated across both ROIs, yielding 200 cells per slide) showed comparable agreement with expert annotations (cross-model MAD 11.4%, with per-model MADs of 11.4% and 11.3% for training with annotations from expert A and B, respectively), whereas inter-expert variability reached an MAD of 11.9%. A non-inferiority analysis showed that model-expert disagreement did not exceed inter-expert disagreement by more than the predefined margin of five percentage points (mean difference = *−*0.54%; one-sided Wilcoxon signed-rank test, *p =* 0.0032). These findings support the use of automated classification for quantitative assessment of local blast frequencies, with deviations comparable to those observed between independent experts.

## Discussion

Diverse computational tasks in hematology—including automated differential counting [9, 15–20], disease classification [21–24, 33–36, 38], and representation learning [29, 32, 48, 49]—share a common prerequisite: the extraction of high-fidelity single-cell patches from raw whole-slide data. Existing approaches, however, are rarely designed for exhaustive whole-slide extraction. Instead, most pipelines rely on multi-stage workflows that restrict computationally demanding full-resolution cell detection to selected regions [9, 16, 21, 23, 26], while publicly available models often show limited transferability to external datasets (Supplementary Table 3). By enabling fast, robust white blood cell localization directly across complete slides without manual intervention, HemAutomaton overcomes these limitations, reducing the need for site-specific retraining or manual adjustments when translating to new laboratory settings.

For practical slide-wide deployment, computational hematology workflows must combine throughput and robustness to variable preparation conditions. NCA models, which are at the core of HemAutomaton, are particularly well suited to these demands. By operating locally, HemAutomaton avoids the heavy parameterization typical of standard vision architectures—such as Faster R-CNN [9], YOLO variants [16, 18, 19], or semantic segmentation networks [21]—which are prone to domain-specific overfitting. Consequently, it matches the in-domain accuracy of Faster R-CNN while maintaining greater robustness under domain shifts, at a reduced computational cost. In addition to computational scalability, annotation efficiency represents a key practical advantage: HemAutomaton achieves near-peak detection performance from as few as 100 annotated cells, substantially lowering the barrier for adapting the model to new datasets.

Large-scale single-cell extraction also provides the necessary data foundation for self-supervised representation learning in computational hematology. Existing curated reference datasets typically range from approximately 28,000 [18] to 171,000 single-cell images [31], and the hematology foundation model DinoBloom was pretrained on a multi-dataset corpus of approximately 380,000 curated white blood cell images [29]. By automatically extracting 108 million single-cell images across the three whole-slide image datasets used in this study, HemAutomaton demonstrates the feasibility of expanding training data volume by nearly three orders of magnitude beyond current curated collections. This scaling capability facilitates the training of broader, more generalizable models [29, 48]. Our few-shot classification experiments demonstrate that pairing pretrained feature representations with sparse target-domain supervision substantially improves cell-type classification over zero-shot transfer, showing that even minimal manual labeling can markedly enhance adaptation to new clinical settings. Together, these findings indicate that HemAutomaton can support both the data curation required to train hematology foundation models and their downstream sample-efficient deployment. Moving beyond single-cell detection and classification, automated whole-slide analysis enables the assessment of macro-spatial patterns of cell-type distributions. In this work, we used HemAutomaton to map the spatial heterogeneity of blast cells within BM and PB smears. While the non-uniform distribution of different cell types across smears—particularly the accumulation of larger cells such as blasts near the feathered edge—is recognized in clinical practice [47], measuring these macroscopic patterns has remained impractical with manual workflows. HemAutomaton overcomes these sampling constraints by profiling cell distributions across entire slides, systematically quantifying the blast gradient and providing an empirical characterization of local sampling biases.

Several limitations of our approach should be acknowledged. First, the macro-spatial analyses rely directly on model predictions and may be subject to localized detection biases; specifically, high cell density in crowded smear regions can disproportionately hinder the detection of larger cells, such as blasts, potentially confounding the observed spatial gradients. Second, the framework involves an inherent trade-off between throughput and image resolution: our lightweight NCA architecture is optimized for exhaustive slide-wide scanning, whereas highly parameterized models or models operating at higher resolution may be preferable when the clinical workflow is restricted to small, pre-selected ROIs. Third, our preprocessing relies on brightness-based heuristics for tile filtering, which may require manual tuning for variable slide backgrounds. Fourth, the current implementation of HemAutomaton focuses on white blood cells exclusively. The detection and analysis of other diagnostically relevant components such as red blood cell morphology, platelet clumping, and smudge cells are beyond the scope of this work and left for future analysis. Fifth, while our three-cohort evaluation validates the framework’s cross-cohort generalizability, its translation into clinical practice requires broader testing across diverse scanners and disease profiles.

In summary, HemAutomaton enables exhaustive single-cell profiling across whole-slide hematology smears. By eliminating manual region selection and requiring minimal annotation for training, the pipeline overcomes conventional sampling constraints to capture complete cellular populations across entire specimens. The parameter-efficient architecture facilitates both large-scale dataset curation for future foundation model training and downstream clinical evaluation, providing a framework for more sensitive AI-supported hematological assessments.

## Methods

### Extraction Pipeline

#### Overview

Single-cell detection is performed using a multi-resolution approach, in which cells are detected at a lower resolution to reduce computational cost. All computations are defined relative to a fixed physical target scale corresponding to the resolution at which the detection model was trained. For each input WSI in a pyramidal format, the effective resolutions of the available pyramid levels are determined, and detection is performed at the pyramid level whose resolution most closely matches the target scale. This strategy avoids unnecessary interpolation while ensuring that the detection model operates at a reasonably consistent physical scale across slides. Non-pyramidal images are rescaled to the target scale.

WSIs or large input images are subdivided into regions with optional overlap, and regions containing little relevant information are excluded prior to inference using brightness heuristics. The NCA-based detection model produces spatial activation maps from which single-cell locations are extracted. When overlap is used, spatially redundant detections are resolved using a distance-based merging criterion. Final detected locations are subsequently mapped back to the full-resolution image. An optional quality filter is included, which is implemented using MobileNetV3 [50] to identify and remove low-quality patches, containing smudged cells, blurry images, and other artifacts.

#### NCA-based cell detection

At the core of HemAutomaton lies an NCA designed for white blood cell center detection. The NCA embeds the RGB image into a latent state and refines this state through repeated local update steps [40]. In our implementation, the model uses a 6-channel state, a hidden size of 128, and a fire rate of 0.5. Each update step applies two learned 3 *×* 3 convolutions, concatenates the resulting local features with the current state, and passes them through a two-layer multilayer perceptron to complete the update step. The RGB channels are reset to the original image content after each update so that the image information remains available while the latent channels evolve.

The NCA is trained from simple cell-center point annotations rather than bounding boxes or segmentations. Point labels are converted into Gaussian heat maps that serve as training targets, and during optimization, a weighted mean squared error is minimized between the predicted response map and the Gaussian target map (Supplementary Figure 6). Training augmentation consists of random horizontal and vertical flips, and the automaton is unrolled for a randomly sampled number of update steps during training. At inference, one latent channel is interpreted as a response map for cell presence. Candidate cell centers are extracted by Gaussian smoothing followed by local-maxima detection and very closely spaced detections are removed by radius-based deduplication.

### Datasets

We assembled data from four independent hematology imaging datasets, comprising whole-slide images (WSIs), manually annotated regions of interest (ROIs), and labeled single-cell images. These data were curated and combined in different configurations for our experiments. An overview of the datasets and their use across this study is provided in the Supplementary Material (Supplementary Figure 1).

#### WSI data

We used three independent whole-slide microscopy datasets comprising blood and bone marrow smears from different institutions and imaging systems (Figure 1A). The B-ALL dataset comprises 240 WSIs, including 120 BM and 120 PB smears in MRXS format, acquired using a 3DHISTECH P1000 scanner. The Höfener et al. dataset comprises 246 BM WSIs in DICOM format, acquired using 3DHISTECH PANNORAMIC MIDI II and PANNORAMIC 250 Flash scanners. The de Almeida et al. dataset comprises 362 cropped PB WSIs in TIFF format, acquired using a Hamamatsu NanoZoomer 2.0 scanner.

#### ROI data

Detection annotations were assembled from the B-ALL, Höfener et al., and de Almeida et al. datasets and harmonized to a common one-class white blood cell detection task. For the B-ALL dataset, manual annotations were generated from 10 patients, each represented by one BM and one PB smear. Two ROIs were selected from each of the 20 smears, yielding 40 ROIs in total. Each ROI was constructed to contain exactly 100 annotated white blood cells, by annotating cells sequentially from left to right within a preselected rectangular region and cropping the ROI immediately after the 100th cell, resulting in 4, 000 annotated cells (Figure 4E). The Höfener et al. dataset comprised 226 manually annotated BM ROIs with 22, 631 WBC annotations after excluding instances labeled as artifact, erythrocyte, giant platelet, megakaryocyte, neutrophil extracellular trap, spicule, thrombocyte, or thrombocyte aggregate. For the de Almeida et al. dataset, we used the AC1 subset comprising 158 image patches from 31 acquisition groups. As AC1 provides segmentation masks rather than point-based detection annotations, binary masks were converted to cell-center annotations by identifying connected components and using their centroids as detection targets. Components smaller than 16 pixels were discarded, yielding 2, 839 annotated cells.

#### Single-cell data

Single-cell annotations were available for the B-ALL, Höfener et al., and Matek et al. datasets. The 4, 000 manually annotated B-ALL cells described above were independently classified by two experts as blast or non-blast cells. The Höfener et al. and Matek et al. datasets provide multiclass annotations for approximately 27, 000 and 171, 000 single-cell images, respectively. As the two datasets use different original annotation schemes, their labels were mapped to a shared set of six morphological categories: blast cells, neutrophils and progenitors, eosinophils/basophils, nucleated erythroid cells, mononuclear cells and progenitors, and other. The complete mapping between the original annotations and the harmonized cell-type scheme is provided in the Supplementary Material (Supplementary Figure 4).

### Three-domain detection benchmark

To evaluate robustness across domains, we established a three-domain benchmark using the curated BALL, Höfener et al., and de Almeida et al. ROI datasets described above. All datasets were evaluated as a common one-class center-detection task. Images were processed at an 8*×* downsampled resolution. Each ROI was converted into overlapping 512 *×* 512 pixel tiles with stride 384, with edge padding to full tile size. Empty tiles were retained with a probability of 0.25 during training, whereas all tiles were retained during validation.

Five-fold cross-validation was performed independently for each source domain, with ROIs originating from the same patient or acquisition group assigned to the same fold. For each fold and source domain, one learned detection model was trained on the source-domain training partition and evaluated both on the held-out in-domain partition and on the two target domains. The same trained checkpoint was reused unchanged for the two cross-domain evaluations from that source domain.

#### Training and baselines

The NCA was trained for 20 epochs with Adam (lr = 10*^−^*^3^, batch size = 2). Point annotations were converted into Gaussian center maps with standard deviation 3.0 pixels and truncation 6.0, and training minimized a weighted mean squared error loss with positive and negative weights of 8.0 and 1.0, respectively. Both detection models were optimized using the same training data augmentation, consisting of random horizontal and vertical flips of the image patches. During training, the NCA was unrolled for 6 to 10 update steps, with the exact number randomly sampled for each batch. At inference time, full-tile response maps were reconstructed from overlapping 256 *×* 256 patches with stride 128. The post-processing threshold was selected on the training split by sweeping thresholds between 0.03 and 0.30 to maximize the F1 score. The last training checkpoint was used for evaluation. We compared HemAutomaton’s NCA-based detection model with Faster R-CNN [46] and a blob baseline evaluated on the same tile datasets. The Faster R-CNN model used a ResNet-50-FPN backbone with random initialization [46, 51]. The model was trained for 20 epochs with stochastic gradient descent (learning rate 5 *×* 10*^−^*^4^, momentum 0.9, weight decay 1 *×* 10*^−^*^4^, and batch size 2). Anchor sizes were set to 8, 16, 32, 64, and 128 pixels with aspect ratios of 0.75, 1.0, and 1.33. Detection thresholds were calibrated on the training split by maximizing center-based F1 scores using a match radius of 8 pixels over thresholds between 0.01 and 0.20. For evaluation, predicted boxes were reduced to their center coordinates and greedily deduplicated within 8 pixels. Because NCA and Faster R-CNN represent different detector families with different parameterizations and loss landscapes, each model was optimized using an optimizer commonly used for that architecture rather than enforcing a shared optimizer across methods.

The classical blob baseline used no learned parameters. Instead, it generated a hand-crafted response map from each RGB tile by combining a purple-favoring chromatic term, grayscale darkness, and an HSV-based mask. The resulting response was Gaussian smoothed (*σ =* 1.2), percentile normalized, and converted to detections by local-maxima extraction. Its absolute response threshold was calibrated on the source-domain training split by maximizing center-based F1 scores.

#### Evaluation metric

All experiments were evaluated as center-detection tasks. Predicted and ground-truth centers were matched one-to-one if their Euclidean distance was within 8 pixels (*∼* 8*µm*); unmatched predictions were counted as false positives and unmatched annotations as false negatives. We regrouped tiles into their parent ROIs and computed F1 scores per ROI. Supplementary Table 1 summarizes the mean and standard deviation across the five folds.

### Computational efficiency evaluation

We benchmarked the computational efficiency of NCA, Faster R-CNN, and the classical blob baseline using an identical tile-based inference pipeline. Experiments were conducted on tiles of size 512 *×* 512 with a physical resolution of approximately 1*µm* per pixel. All measurements were performed on a system equipped with an Intel Xeon Gold 6226R CPU and an NVIDIA Tesla V100S GPU (32 GB VRAM). Benchmarks were executed using Python 3.10 with PyTorch and CUDA support. For each method, inference was evaluated on the same set of 12 tiles with batch size 1. Two warm-up passes were performed before timing, followed by two repeated measurements per tile. To ensure consistent measurements, CPU threading was restricted to a single thread. We reported the number of model parameters, floating-point operations (FLOPs) per tile, latency per tile, throughput in tiles per second, and peak memory usage during inference.

### Low-data generalization experiment

To quantify sample efficiency under increasing domain shifts, we conducted a low-data experiment where we fixed the training domain to B-ALL BM and systematically varied the amount of single-cell training data. Using the grouped five-fold split from the detection benchmark, we constructed nested budgets of *k =* 1, …, 8 source slides. For each selected slide, one ROI was retained for training, so that each budget increment contributed one additional source slide and no two selected ROIs originated from the same slide.

To standardize annotation volume and eliminate intra-slide sampling bias, each training patch was extracted from a distinct source slide and contained exactly 100 annotated white blood cells. Specifically, cells were annotated sequentially from left to right within a pre-selected rectangular ROI, with the patch cropped immediately following the 100th annotated white blood cell. Consequently, each incremental budget step contributed exactly 100 additional annotated cells originating from a unique slide, ensuring no two patches shared a common slide origin. For each budget and fold, NCA and Faster R-CNN were trained from scratch with the same model configurations, augmentation scheme, optimization settings, and threshold-selection procedures used in the full detection benchmark. The blob detection baseline was excluded from this experiment, as the full-data benchmark had already established its inability to achieve generalizable cell localization.

Trained models were evaluated across four distinct validation scenarios: (i) held-out B-ALL BM (in-domain), (ii) B-ALL PB from the same cohort (cross-modality), (iii) the independent BM smear dataset from Höfener et al. [18] (cross-center), and (iv) the independent PB smear dataset from de Almeida et al. [21] (combined cross-modality and cross-center shift). Detection performance was quantified using F1 scores and reported as mean and standard deviation across the five validation folds.

### Few-shot classification

Few-shot classification was evaluated using the curated and harmonized Matek et al. and Höfener et al. single-cell datasets described above. Single-cell images were embedded using the frozen pretrained DinoBloom-S feature extractor. To obtain a morphologically diverse set of target-domain cells for annotation, embeddings were clustered using k-means and representative cells were selected from each cluster.

#### Prototype- and example-based classification

Following annotation of the selected examples, the remaining target cells were classified using cosine similarity in the DinoBloom embedding space. For each query cell, the five nearest annotated target cells were identified and their labels combined by majority vote. To account for cell types that are absent or only sparsely represented among the labeled target cells, source-domain prototypes were additionally incorporated. Source-domain prototypes were computed by averaging feature embeddings within each fine-grained source class and subsequently mapped to the harmonized target label space. Source- and target-derived class probabilities were combined according to

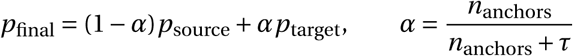

where *n*_anchors_ denotes the number of labeled target examples available for the corresponding harmonized cell type and *τ* is a regularization parameter. As additional target annotations become available, the prediction progressively shifts from source-domain prototypes towards target-domain examples.

#### Experimental setup

For each target dataset, five folds were generated, where one fold was held out for testing while the remaining cells formed the target pool for cluster-guided exemplar selection. The number of clusters was set to the number of fine-grained cell categories present in the respective dataset, and we set *τ* to 25. We evaluated annotation budgets of *k ∈* {1, 2, 5, 10, 20} representative cells per cluster. Performance was summarized using macro-F1 scores. Supplementary Table 4 reports results for all annotation budgets, while Supplementary Figure 5 shows the confusion matrices of the best-performing configurations.

### Spatial variance of blast frequencies

Spatial analyses were performed on the B-ALL dataset using single-cell images automatically extracted by the proposed pipeline. For the descriptive whole-slide analyses, the few-shot classifier was adapted to binary blast versus non-blast classification using all 4, 000 expert-annotated single-cell images from the B-ALL dataset. Evaluation of the classifier for blast-frequency estimation was performed separately using patient-level cross-validation.

#### Local blast frequency

For each detected cell, the local blast frequency was estimated from its 500 nearest neighboring cells. The local blast frequency was defined as the fraction of cells predicted as blasts within this neighborhood. In this way, each detected cell serves as a discrete sampling point that represents the local cellular composition.

#### Smear position

To quantify spatial trends along the smear, each detected cell was assigned a relative position within the analyzable smear region. Tiles passing the brightness filter were grouped into contiguous vertical smear segments. For each segment, the mean grayscale intensity of the uppermost and lowermost three tiles was compared to identify the bright (feathered edge) and dark (dense smear) ends. The relative smear position of cells was defined as the normalized distance between these two ends, yielding values between 0 (dense smear region) and 1 (feathered edge). Cells that could not be assigned to a valid smear segment were excluded from smear-position analyses.

#### Blast-frequency variability and model agreement

To assess the reliability of local blast-frequency estimates, we used the 4, 000 manually annotated cells from 20 B-ALL slides (10 patients with one BM and one PB slide each). Two independent hematology experts (A and B) classified the same 100 cells in each of two ROIs per slide as blast or non-blast cells. Blast frequencies were computed independently for each ROI and expert.

Spatial sampling variability was quantified with MAD by comparing blast frequencies between the two ROIs of each slide, while inter-expert variability was assessed by comparing the two experts’ estimates for the same aggregated ROIs (2 ROIs = 200 cells per slide).

To evaluate automated blast-frequency estimation, two binary classifiers were independently trained using annotations from expert A or expert B in a 10-fold patient-level cross-validation. Models were evaluated using slide-level grouped cross-validation to prevent cells from the same slide appearing in both training and test sets. Predictions from each model were compared against the independent expert annotations (model A versus expert B and model B versus expert A).

For each slide *i*, inter-expert disagreement was defined as

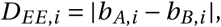

where *b_A_*,*_i_* and *b_B_*,*_i_* denote the blast frequencies estimated by experts A and B, respectively. Cross-expert model disagreement was defined as

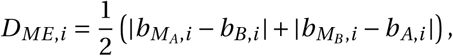

where *b_M__A_*, *i* and *b_M__B_*, *i* are the corresponding model-derived blast frequencies. The difference in disagreement was

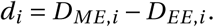

Non-inferiority was assessed using a margin of *δ =* 0.05. A one-sided Wilcoxon signed-rank test was applied to *d_i_ −δ* across the 20 slides, testing whether the location of the paired differences was below the specified margin. Statistical significance was defined as *p <* 0.05.

## Data availability

Publicly available WSI smear and single-cell data used in this work were described in the original studies [18, 21, 31] and accessed via the Cancer Imaging Archive (https://www.cancerimagingarchive.net/collection/bone-marrow-cytomorphology_mll_helmholtz_fraunhofer/), Zenodo (https://zenodo.org/records/17486627), and the BioImage Archive (https://www.ebi.ac.uk/biostudies/BioImages/studies/S-BIAD440).

The B-ALL dataset is not publicly available due to data protection regulations. However, these data may be shared upon reasonable request, subject to institutional and ethical approvals and the execution of a formal data transfer agreement. This research has been approved by the ethics committee of the Friedrich-Alexander University of Erlangen-Nuremberg (Application 22-148-Br).

## Code availability

Code for data preprocessing, model architectures, and model optimization, as well as optimized model weights will be made publicly available upon publication.

## Acknowledgements

C. Marr received funding from the European Research Council under the European Union’s Horizon 2020 Research and Innovation Programme (grant agreements 866411, 101113551, and 101213822) and support from the Hightech Agenda Bayern. This work was supported through the Bavarian AI Foundation Model Initiative, funded by the Bavarian State Ministry of Science and the Arts (StMWK). M. Metzler received a research grant from “Schornsteinfeger helfen krebskranken Kindern e.V.” (Dörfles-Esbach, Germany). The authors thank Prof. Dr. Gunnar Cario and Birthe Fedders (University Hospital Schleswig-Holstein) for supplying the PB and BM smears. The authors thank Stefanie Barnickel, Nathalie Dollmann, and Tatjana Flamann for technical support with cell classification.

## Supplementary Material

**Supplementary Figure 1:**
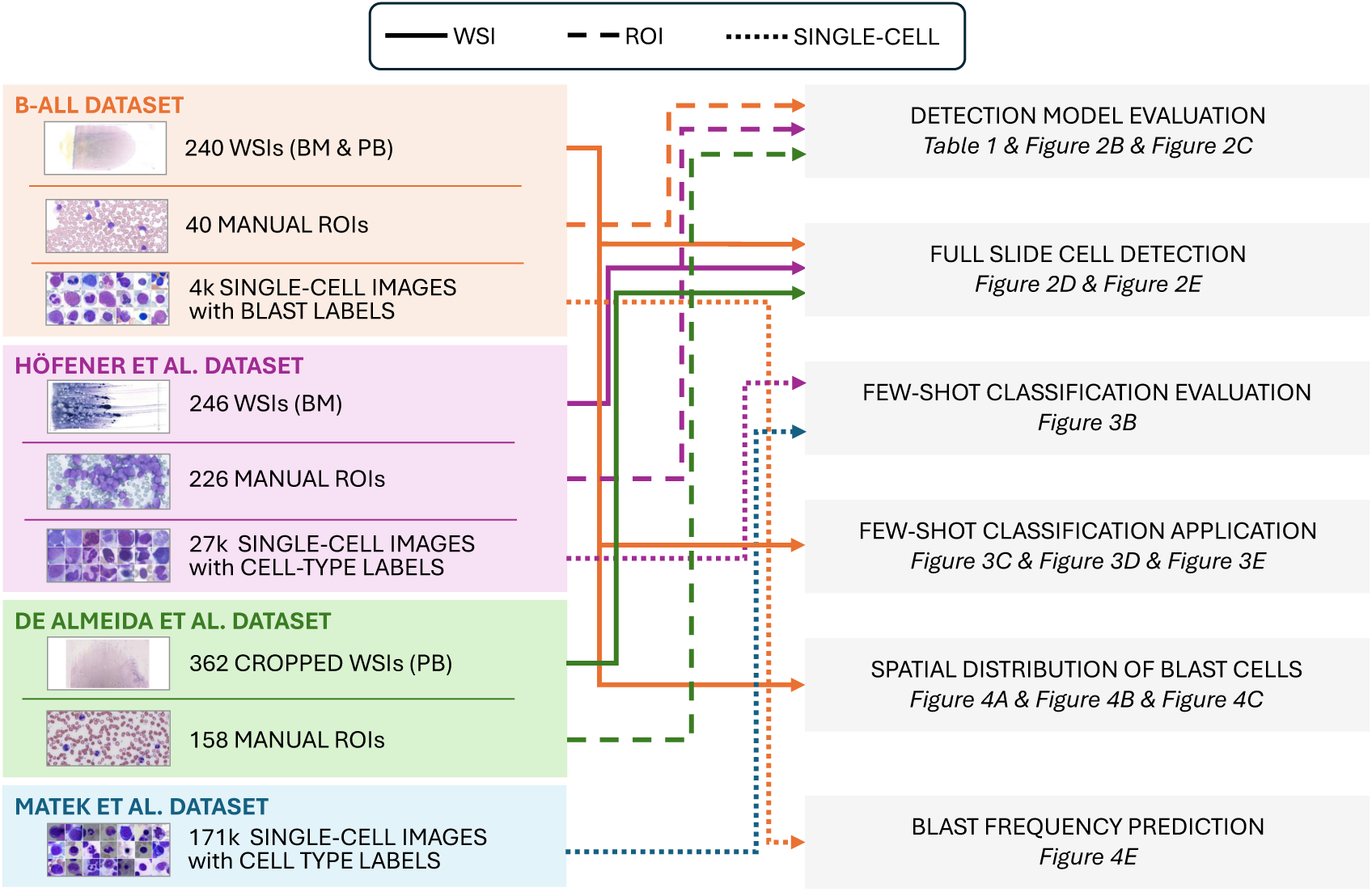
Four hematology datasets provide image data and annotations for experiments throughout the study. The B-ALL dataset contains BM and PB WSIs, manually annotated ROIs with cell coordinates, and blast-level single-cell annotations. The Höfener et al. dataset provides BM WSIs, manually annotated ROIs with bounding boxes, and labeled single-cell images. The de Almeida et al. dataset contains coarsely cropped PB WSIs and manually annotated ROIs with segmentation masks. The Matek et al. dataset provides labeled single-cell images only. Solid, dashed, and dotted arrows indicate the use of WSIs, ROI patches, and single-cell images, respectively.

**Supplementary Table 1:** NCA shows consistent white blood cell detection performance across in-domain and cross-domain evaluation. Results are shown for the complete train-test matrix across the B-ALL, Höfener et al., and de Almeida et al. datasets. Blob, Faster R-CNN, and NCA are trained using one dataset at a time and evaluated on all three datasets. ROI-level macro-F1 scores (mean ± standard deviation) are obtained from five-fold cross-validation. The best-performing method for each train-test combination is highlighted in bold. Table 1 summarizes these results by averaging the three in-domain and six cross-domain train-test combinations for each detection method.

| Trained on | Tested on | Blob | Faster R-CNN | NCA |
| --- | --- | --- | --- | --- |
| B-ALL dataset | B-ALL dataset | $0.50 \pm 0.11$ | $0.83 \pm 0.06$ | **$0.86 \pm 0.04$** |
| | Höfener et al. | $0.49 \pm 0.07$ | $0.63 \pm 0.15$ | **$0.84 \pm 0.01$** |
| | de Almeida et al. | $0.27 \pm 0.12$ | $0.63 \pm 0.21$ | **$0.90 \pm 0.03$** |
| Höfener et al. | B-ALL dataset | $0.54 \pm 0.01$ | **$0.82 \pm 0.02$** | $0.81 \pm 0.01$ |
| | Höfener et al. | $0.82 \pm 0.02$ | **$0.88 \pm 0.01$** | $0.84 \pm 0.01$ |
| | de Almeida et al. | $0.34 \pm 0.24$ | $0.84 \pm 0.06$ | **$0.89 \pm 0.03$** |
| de Almeida et al. | B-ALL dataset | $0.61 \pm 0.11$ | **$0.70 \pm 0.09$** | $0.59 \pm 0.25$ |
| | Höfener et al. | **$0.80 \pm 0.01$** | $0.29 \pm 0.14$ | $0.71 \pm 0.05$ |
| | de Almeida et al. | $0.45 \pm 0.12$ | **$0.89 \pm 0.03$** | $0.85 \pm 0.08$ |

**Supplementary Figure 2:**
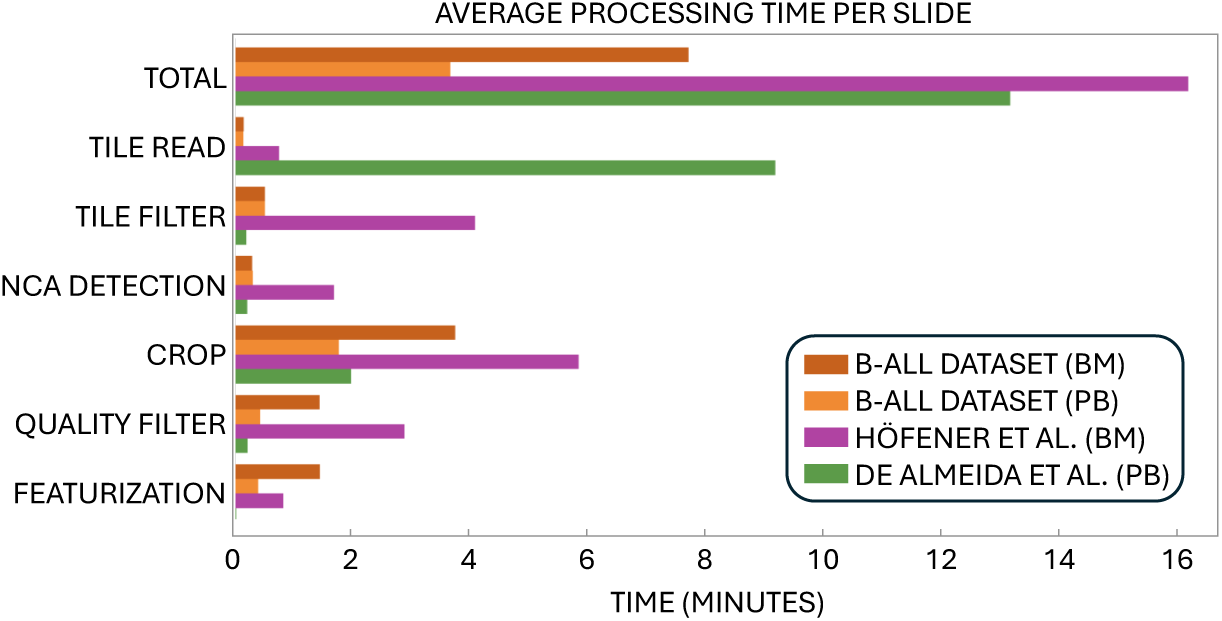
Full-slide processing of up to hundreds of thousands of cells is completed within minutes. Average processing time per slide is shown for the B-ALL bone marrow (BM), B-ALL peripheral blood (PB), Höfener et al. (BM), and de Almeida et al. (PB) datasets, separated into individual processing steps from data reading to single-cell feature extraction. Total processing time varies across datasets, reflecting differences in slide format, size, and cellular content. Processing time for the de Almeida et al. dataset is dominated by tile reading as these WSIs are provided at a single resolution and therefore require on-the-fly rescaling. On the Höfener et al. dataset, tile reading, filtering, and detection take longer relative to the other datasets because the available pyramid levels are more coarsely spaced and the pipeline chooses the next higher-resolution pyramid level relative to its target resolution, resulting in more tiles to be processed.

**Supplementary Table 2:** Only a subset of published whole-slide white blood cell detection pipelines provides the resources required for inference. Studies were assessed for the availability of public source code and pretrained model weights required to reproduce WBC detection on external whole-slide images. Tayebi et al., de Almeida et al., and Kockwelp et al. provided sufficiently documented implementations and pretrained weights and were therefore included in the experimental evaluation. Lewis et al. provided pretrained model files but no documented executable pipeline enabling reliable reproduction of the complete workflow. Wang et al. [25] released code for their benchmarking framework but referred to Cellpose-SAM [52] for cell segmentation, without providing the additional filtering or classification step required to distinguish white blood cells from other segmented objects. Commercial systems and studies for which no public implementation was identified were not included.

| Study | Modality | Code | Weights | Note |
| --- | --- | --- | --- | --- |
| *Not included in experimental evaluation* |  |  |  |  |
| Fu et al., 2020 [15] | BM | × | × | Commercial system |
| Lv et al., 2023 [17] | BM | × | × | Commercial system |
| Höfener et al., 2025 [18] | BM | × | × | No public code statement |
| Wang et al., 2022 [22] | BM | × | × | No public code statement |
| Sun et al., 2025 [19] | BM | × | × | No public code statement |
| Zhou et al., 2025 [20] | BM | × | × | No public code statement |
| Lewis et al., 2023 [9] | BM | × | ✓ | No documented pipeline |
| Yan et al., 2026 [24] | BM | ✓ | × | Weights release pending |
| Wang et al., 2026 [25] | PB | Partial | Partial | Refers to Cellpose-SAM |
| *Included in experimental evaluation* |  |  |  |  |
| Tayebi et al., 2022 [16] | BM | ✓ | ✓ | Full pipeline available |
| de Almeida et al., 2023 [21] | PB | ✓ | ✓ | Full pipeline available |
| Kockwelp et al., 2023 [23] | BM | ✓ | ✓ | Full pipeline available |

**Supplementary Table 3:** NCA achieves the highest average cross-dataset white blood cell detection performance across three independent WSI datasets compared to published models. F1 scores were evaluated on annotated ROIs from three independent WSI datasets. Publicly available pretrained models were evaluated without changes. For NCA, performance on each dataset represents the average of the two cross-domain experiments in which the model was trained on either of the other two datasets. The average denotes the mean F1 score across the three evaluation datasets. The best-performing method for each dataset and on average is highlighted in bold.

| Method | B-ALL dataset | Höfener et al. | de Almeida et al. | Average |
| --- | --- | --- | --- | --- |
| Kockwelp et al. | 0.00 | 0.01 | 0.63 | 0.21 |
| de Almeida et al. | 0.01 | 0.16 | 0.86 | 0.34 |
| Tayebi et al. | 0.22 | **0.81** | 0.32 | 0.45 |
| **NCA** | **0.70** | 0.76 | **0.90** | **0.79** |

**Supplementary Figure 3:**
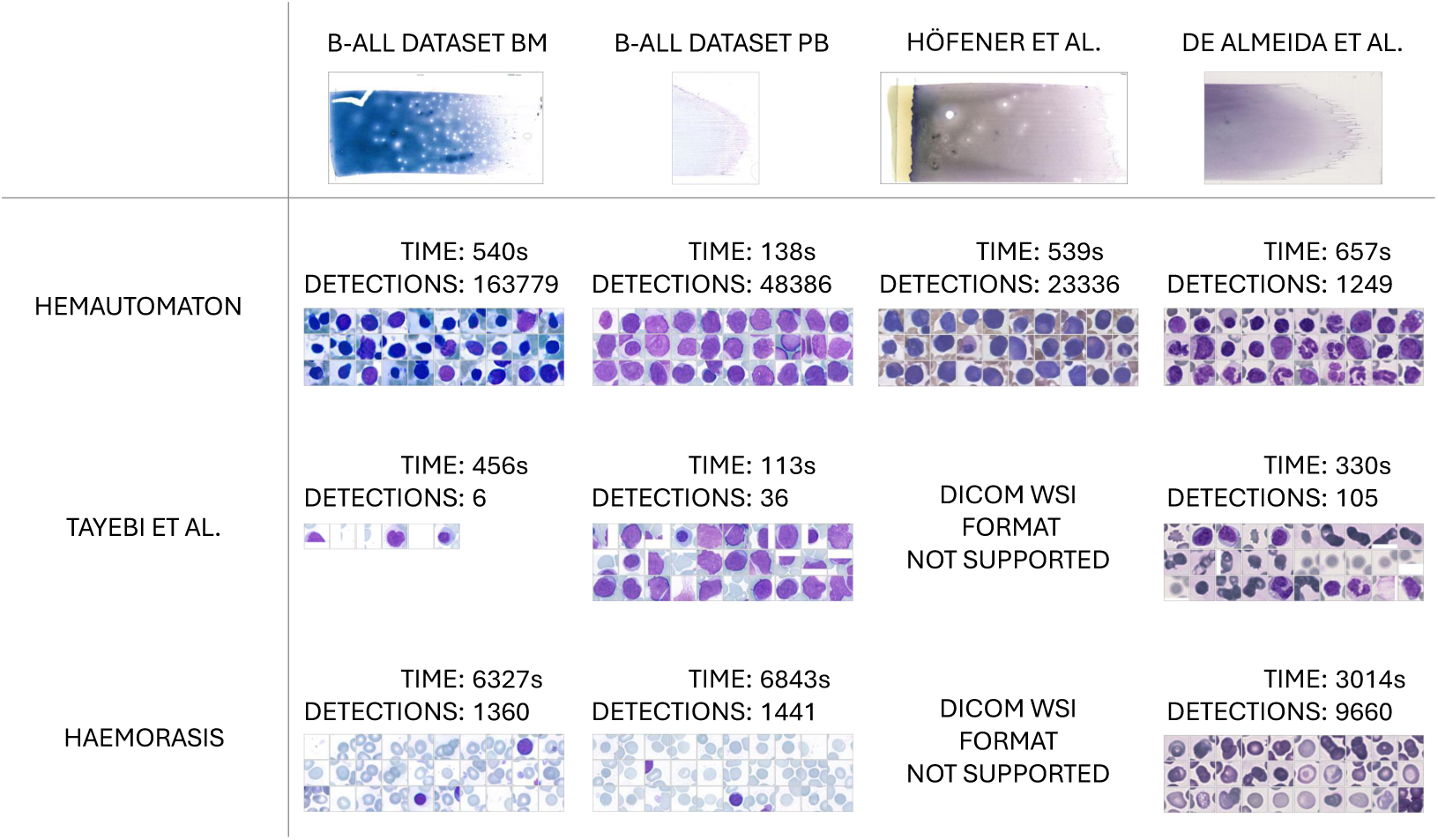
Published WBC detection pipelines show limited transferability when deployed on external whole-slide images without dataset-specific adaptation. HemAutomaton and the publicly released pipelines of Tayebi et al. and de Almeida et al. were applied without modification to one representative WSI from each supported dataset. Processing time and total number of detections are reported for each pipeline, together with 30 randomly selected detections shown as single-cell images. The Tayebi et al. pipeline discarded more than 99% of tiles during preprocessing and consequently processed the slides rapidly, but yielded less than 150 detections across slides. The de Almeida et al. pipeline produced more detections but required substantially longer processing times than HemAutomaton and showed frequent non-WBC detections among the randomly sampled cells. Neither public pipeline supported DICOM format and was therefore not applied to the Höfener et al. slide. The Kockwelp et al. method, while included in the ROI-level benchmark, was not evaluated at the slide level because its released implementation required an incompatible input data format. Because reproducing third-party pipelines is susceptible to user error and may depend on implementation details not fully captured by the available documentation, these results reflect our best-effort application of the publicly released implementations and should not be interpreted as definitive estimates of their performance.

**Supplementary Table 4:**
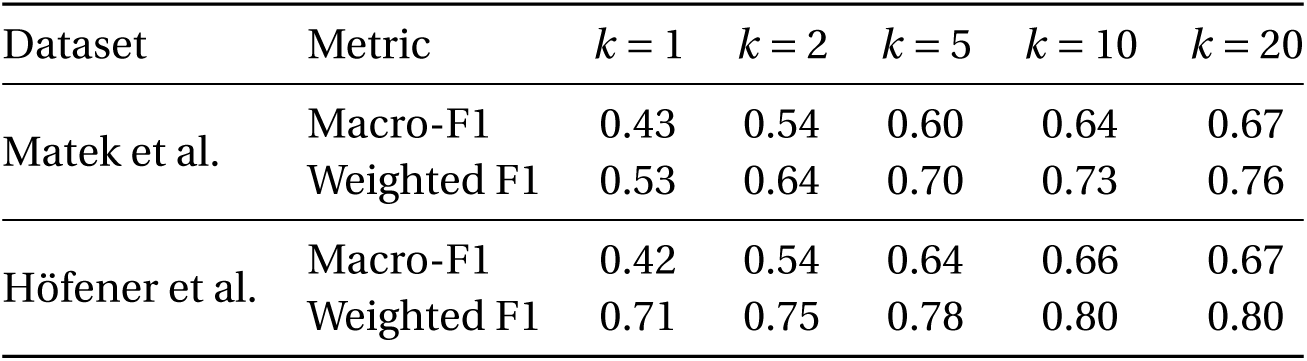
Few-shot classification performance improves consistently with increasing annotation budget. Classification performance for different numbers of manually labeled target-domain cells per cluster (k) is reported as macro-F1 and weighted-F1 on the Matek et al. and Höfener et al. datasets. The number of clusters was set to the number of original fine-grained cell-type labels in each dataset, resulting in 21 and 48 clusters for the Matek et al. dataset and Höfener et al. dataset, respectively.

**Supplementary Figure 4:**
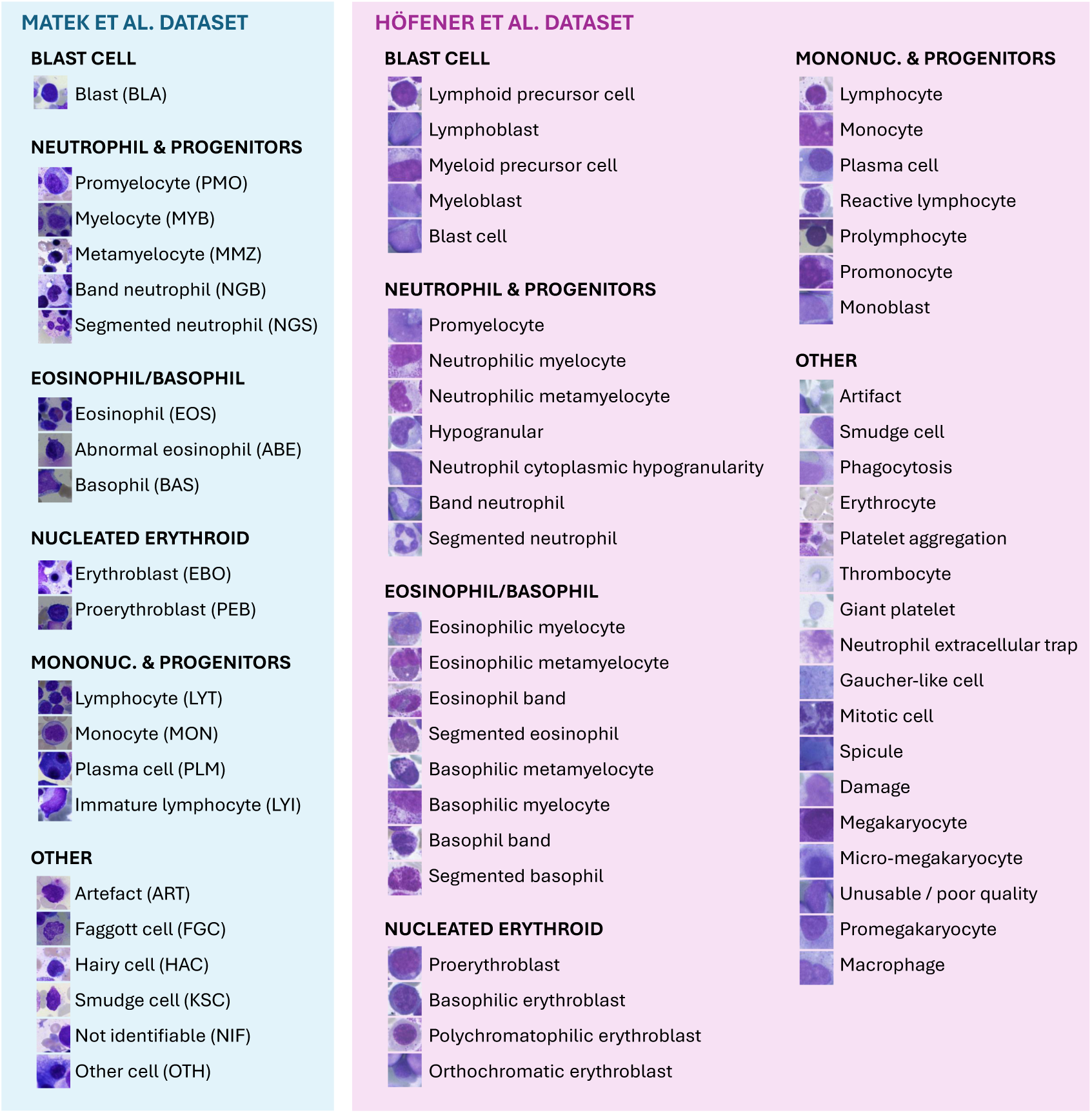
Harmonization of fine-grained cell-type annotations allows for consistent cross-dataset evaluation. Fine-grained labels from Matek et al. and Höfener et al. are mapped to a shared set of six harmonized morphological categories. Each category shows representative single-cell images together with the corresponding fine-grained labels. Abbreviations shown for the Matek et al. dataset correspond to those used in the original dataset.

**Supplementary Figure 5:**
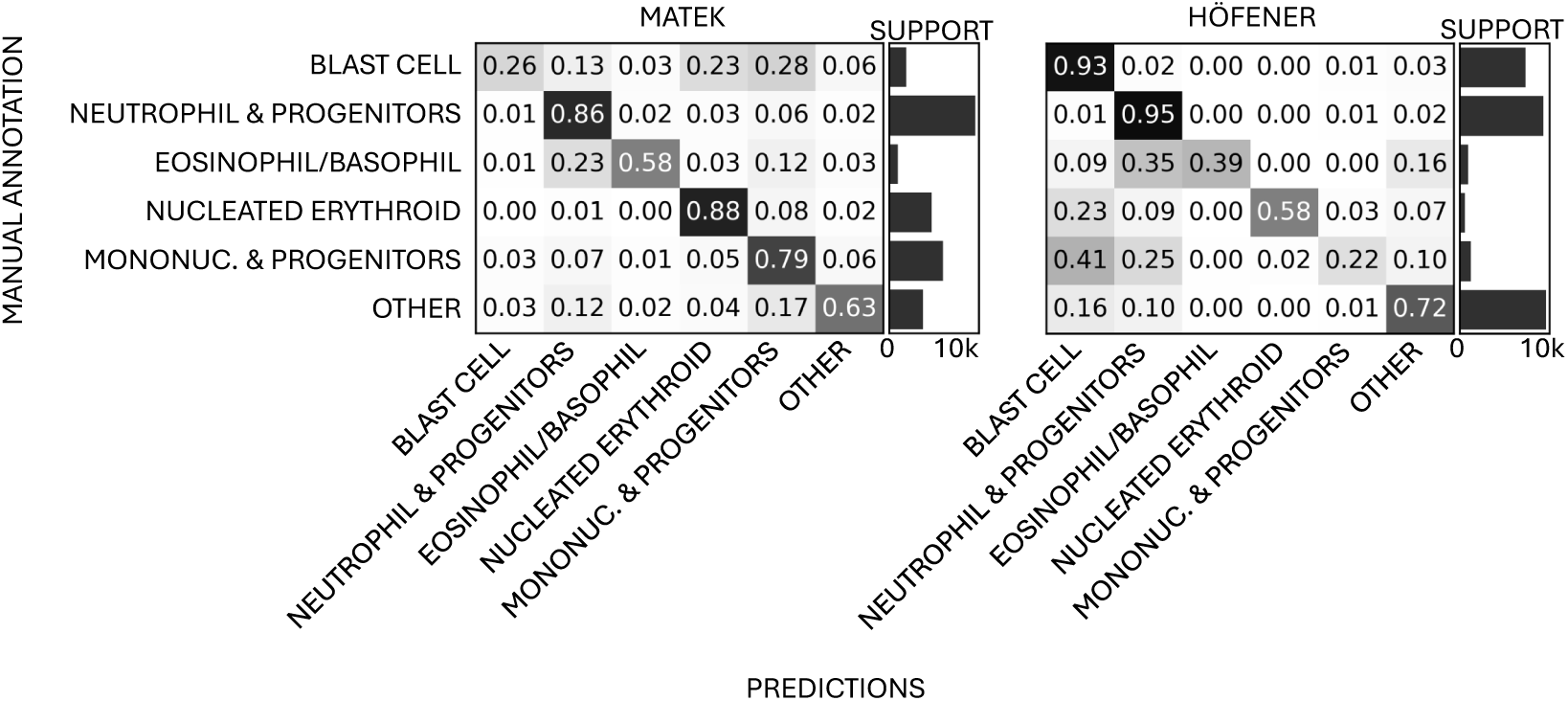
Few-shot classification identifies well-represented cell types with minimal annotation cost. Normalized confusion matrices show few-shot classification performance on the Matek et al. and Höfener et al. datasets. Predictions were obtained using 20 labeled examples per cluster. Rows indicate ground truth labels and columns indicate predictions. Bar plots indicate the support of the evaluation dataset. Frequently represented classes show high accuracy, whereas lower performance is predominantly observed for classes with fewer samples.

**Supplementary Figure 6:**
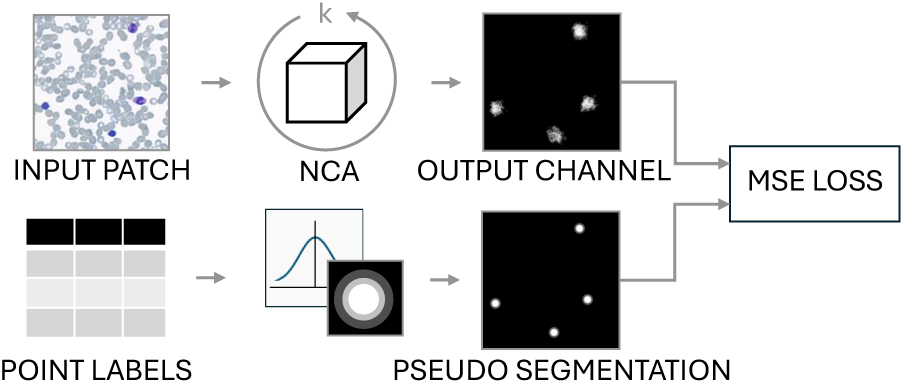
Point annotations provide supervision for the NCA-based white blood cell detection model. During training, the NCA receives an image patch and iteratively updates its internal state to produce a single-channel activation map. Point annotations are converted into pseudo-segmentation targets by applying Gaussian kernels centered at each annotated cell. Model parameters are optimized by minimizing the mean squared error (MSE) between the predicted activation map and the pseudo-segmentation target.

